# Noninvasive Quantification of Radiation-Induced Lung Injury using a Targeted Molecular Imaging Probe

**DOI:** 10.1101/2023.09.25.23295897

**Authors:** Eric Abston, Iris Y. Zhou, Jonathan A. Saenger, Sergey Shuvaev, Eman Akam, Shadi A. Esfahani, Lida P. Hariri, Nicholas J. Rotile, Elizabeth Crowley, Sydney B. Montesi, Valerie Humblet, Grae Arabasz, Ciprian Catana, Florian J. Fintelmann, Peter Caravan, Michael Lanuti

## Abstract

**Rationale:** Radiation-induced lung injury (RILI) is a progressive inflammatory process commonly seen following irradiation for lung cancer. The disease can be insidious, often characterized by acute pneumonitis followed by chronic fibrosis with significant associated morbidity. No therapies are approved for RILI, and accurate disease quantification is a major barrier to improved management.

**Objective:** To noninvasively quantify RILI, utilizing a molecular imaging probe that specifically targets type 1 collagen in mouse models and patients with confirmed RILI.

**Methods:** Using a murine model of lung radiation, mice were imaged with EP-3533, a type 1 collagen probe to characterize the development of RILI and to assess disease mitigation following losartan treatment. The human analog probe targeted against type 1 collagen, ^68^Ga-CBP8, was tested on excised human lung tissue containing RILI and quantified via autoradiography. Finally, ^68^Ga-CBP8 PET was used to assess RILI *in vivo* in six human subjects.

**Results:** Murine models demonstrated that probe signal correlated with progressive RILI severity over six-months. The probe was sensitive to mitigation of RILI by losartan. Excised human lung tissue with RILI had increased binding vs unirradiated control tissue and ^68^Ga-CBP8 uptake correlated with collagen proportional area. Human imaging revealed significant ^68^Ga-CBP8 uptake in areas of RILI and minimal background uptake.

**Conclusions:** These findings support the ability of a molecular imaging probe targeted at type 1 collagen to detect RILI in preclinical models and human disease, suggesting a role for targeted molecular imaging of collagen in the assessment of RILI.

Clinical trial registered with www.clinicaltrials.gov (NCT04485286, NCT03535545)

## Introduction

Lung cancer is among the most common cancer types in the world with 2.2 million new cases annually and a leading cause of cancer death^1^. Radiation therapy is a mainstay of treatment for medically inoperable early-stage lung cancers. Radiation-Induced Lung Injury (RILI) is a side effect of radiotherapy marked by radiographic changes outside of the target region observed in up to 70% of patients^2, 3^. While most patients receiving thoracic radiation develop radiographic changes on chest CT, only a subset of these patients will go on to develop clinically significant RILI. RILI comprises a spectrum of lung disease ranging from acute pneumonitis to chronic fibrosis and is often slowly progressive, with onset of symptoms occurring months after irradiation^4, 5^.

Symptoms commonly include progressive shortness of breath and cough which can be debilitating. Severe cases require supplemental oxygen and sometimes mechanical ventilation. While the study of survival in patients with RILI is complicated by coexisting malignancy, RILI appears to negatively affect survival in small studies^6, 7, 8^.

There are currently no reliable methods to diagnose RILI, predict disease course, or measure treatment effects. RILI remains a diagnosis of exclusion in patients with a history of radiation exposure after other causes of lung disease such as recurrent malignancy and infection have been ruled out. Common grading systems used to quantify RILI rely heavily on individual patient symptoms and lack objective standards^9, 10, 11^. Pulmonary function tests have poor sensitivity, likely due to the relatively small volume of lung typically affected by RILI. High-resolution computed tomography (HRCT) is the gold standard modality used in the clinical evaluation of RILI, but changes in attenuation do not always indicate the presence of disease. ¹⁸F-Fluorodeoxyglucose positron emission tomography (PET), magnetic resonance imaging (MRI), and single photon emission computed tomography have also been used experimentally with some success^12, 13, 14, 15^ but no imaging modality has yet been approved to predict clinically significant RILI or demonstrate response to therapy.

Clinical management of RILI has proven difficult and there is currently no FDA-approved treatment for RILI. Steroids are empirically given to treat pneumonitis, and early studies suggest some benefit with pentoxyfiline and ACE inhibitors in humans and animals^16, 17, 18^. A recent trial evaluating nintedanib in RILI failed to show benefit^19^. The most successful strategies for RILI have come with advances in limiting the radiation dose and volume of irradiated lung with techniques such as stereotactic ablative body radiotherapy (SABR) and intensity modulated radiation therapy (IMRT), as well as through careful risk stratification of patients with known factors such as age, concomitant pulmonary fibrosis, and chemotherapy making RILI more likely^5^.

Collagen is a principal building block used in the creation of fibrotic tissue and is routinely assayed in experimental models of RILI either biochemically or histologically. Pulmonary fibrosis due to RILI becomes apparent on HRCT when high concentrations of collagen and extracellular matrix proteins accumulate in diseased lung tissue.

However, by the time changes are seen on HRCT injury to the lung is likely irreversible. We hypothesize that a more sensitive method to detect increasing deposition of collagen in the lung could identify RILI at an early stage and be useful to identify patients who might benefit from early intervention with antifibrotic therapy.

Our lab has developed molecular imaging probes targeted to type 1 collagen for detecting and staging tissue fibrosis. These probes are based on a 16 amino acid cyclic peptide derived from a phage display screen. An MRI probe termed EP-3533 had 3 gadolinium chelates conjugated to the peptide,^20, 21^ while a PET probe (^68^Ga-CBP8) conjugated the positron emitting isotope gallium-68 for detection. We have demonstrated that EP-3533 enhanced MRI can quantify pulmonary fibrosis in the mouse bleomycin injury model^22^, as well as in rodent models of cardiac, renal, and liver fibrosis^20, 22, 23, 24, 25, 26^. ^68^Ga-CBP8 PET was shown to be specific for pulmonary fibrosis in mouse models and explanted human lung tissue, and that PET signal increased with lung collagen content^27^. In human subjects, ^68^Ga-CBP8 is well tolerated and shows rapid clearance via the kidneys with low signal in healthy lungs^28^. Patients with idiopathic pulmonary fibrosis showed significantly higher lung uptake of ^68^Ga-CBP8.

In this study, we test the hypothesis that fibrosis due to RILI can be identified by type 1 collagen molecular imaging. We developed a murine model of RILI and tested the ability of EP-3533 enhanced MRI to quantify pulmonary fibrosis in RILI over a time course of disease progression, and mitigation treatment with losartan. This common anti-hypertensive medication has known antifibrotic activity and has been successfully tested in radiation exposure mitigation models^29^. To further translate this work, we assayed the specific binding of ^68^Ga-CBP8 to explanted human RILI lung tissue. Finally, we tested the ability of ^68^Ga-CBP8 PET to detect RILI *in vivo* in human subjects with established RILI.

## Results

### Type 1 Collagen Probe Sensitivity to Progression of RILI

We first sought to establish a murine model of RILI, which accurately reproduced its characteristic progressive fibrotic changes, and test whether a type 1 collagen probe could noninvasively measure time-dependent changes in disease severity. Following a single exposure of 21 Gy to the right hemithorax, mice underwent imaging at either 3- month (3M) or 6-month (6M) time points and probe uptake was compared to unirradiated controls (0 Gy) (Figure 1A).

**Figure 1:**
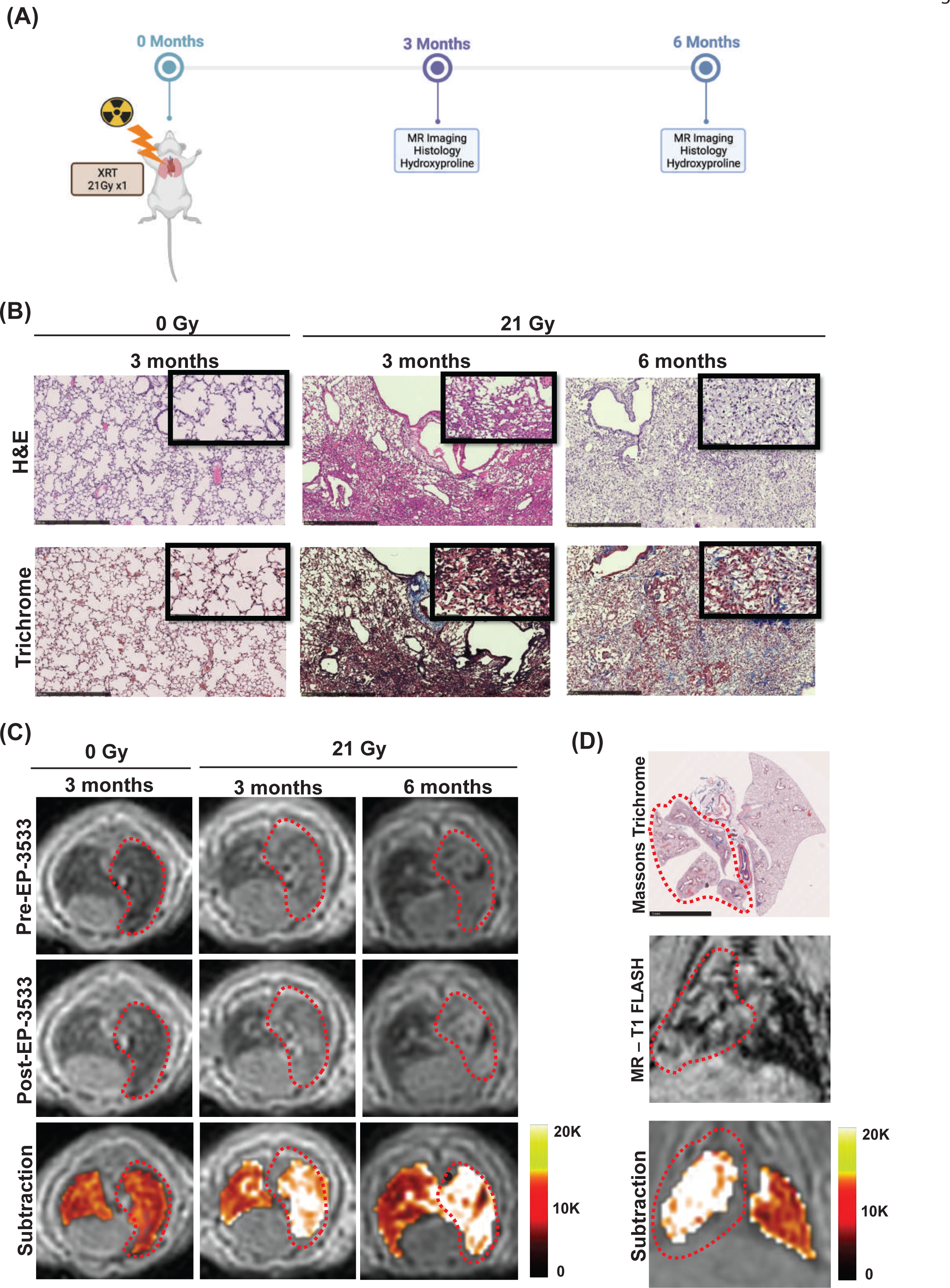
Murine model of progressive RILI. Mice underwent a single dose of 21 Gy to the right hemithorax and were imaged at either 3- or 6-months post irradiation (A). Representative H&E (top) and trichrome (bottom) stains of right lung (B) show normal lung architecture in unirradiated 0M mice. Mice at 3 months developed cellular infiltrate, alveolar thickening, consolidation, and collagen deposition. By 6 months, significant fibrosis and alveolar consolidation was seen. Representative axial MR images (C) are shown before and after EP-3533 injection. Lungs of unirradiated mice (0Gy) show low signal on UTE MR. Irradiated mice show increasing right lung signal from 3 to 6 months due to consolidation, and these areas are enhanced following EP-3533 administration. False color subtraction images (40 minutes following EP-3533 injection – preinjection) overlaid on pre-injection UTE images demonstrate increased signal enhancement in irradiated lung. Co-localization of lung injury in the irradiated right hemithorax on pathology and coronal MR images (D). Diffuse injury throughout the right lung but not in the left lung is seen on Masson’s trichrome and MR-T1FLASH image. False color subtraction image illustrates substantial signal enhancement in the right lung but not in the non-irradiated left lung.

The lungs of irradiated mice demonstrated significant markers of RILI, which were progressive over time. H&E stain and Masson’s trichrome stain demonstrated cellular infiltrate, alveolar thickening, consolidation, and collagen deposition consistent with prior descriptions of RILI^4, 36^, which progressed from 3 to 6-month time points with additional fibrosis and alveolar consolidation (Figure 1B). Collagen proportional area (CPA) was significantly increased in mice at both 3M and 6M time points over nonirradiated controls (ANOVA p<0.001, *post hoc*: 0M 16 ± 0.2% vs 3M 29 ± 0.7%, p<0.001; 0M vs 6M 36 ± 1.9%, p<0.001; 3M vs 6M p=0.03) (Figure 2E). The Ashcroft score was significantly elevated in the irradiated groups as well (ANOVA p<0.001 *post hoc*: 0M 0.06 ± 0.01 vs 3M 2.50 ± 0.15; p<0.001; 0M vs 6M 3.1 ± 0.14, p<0.001, 3M vs 6M p<0.001) (Figure 2F). Hydroxyproline, a measure of collagen deposition in RILI, was also significantly elevated with irradiation (ANOVA p<0.001 *post hoc*: 0M 130 ± 7 µg/lung vs 3M 201 ± 10 µg/lung, p<0.001; 0M vs 6M 260 ± 7 µg/lung p<0.001; 3M vs 6M p=0.003) (Figure 2D).

**Figure 2:**
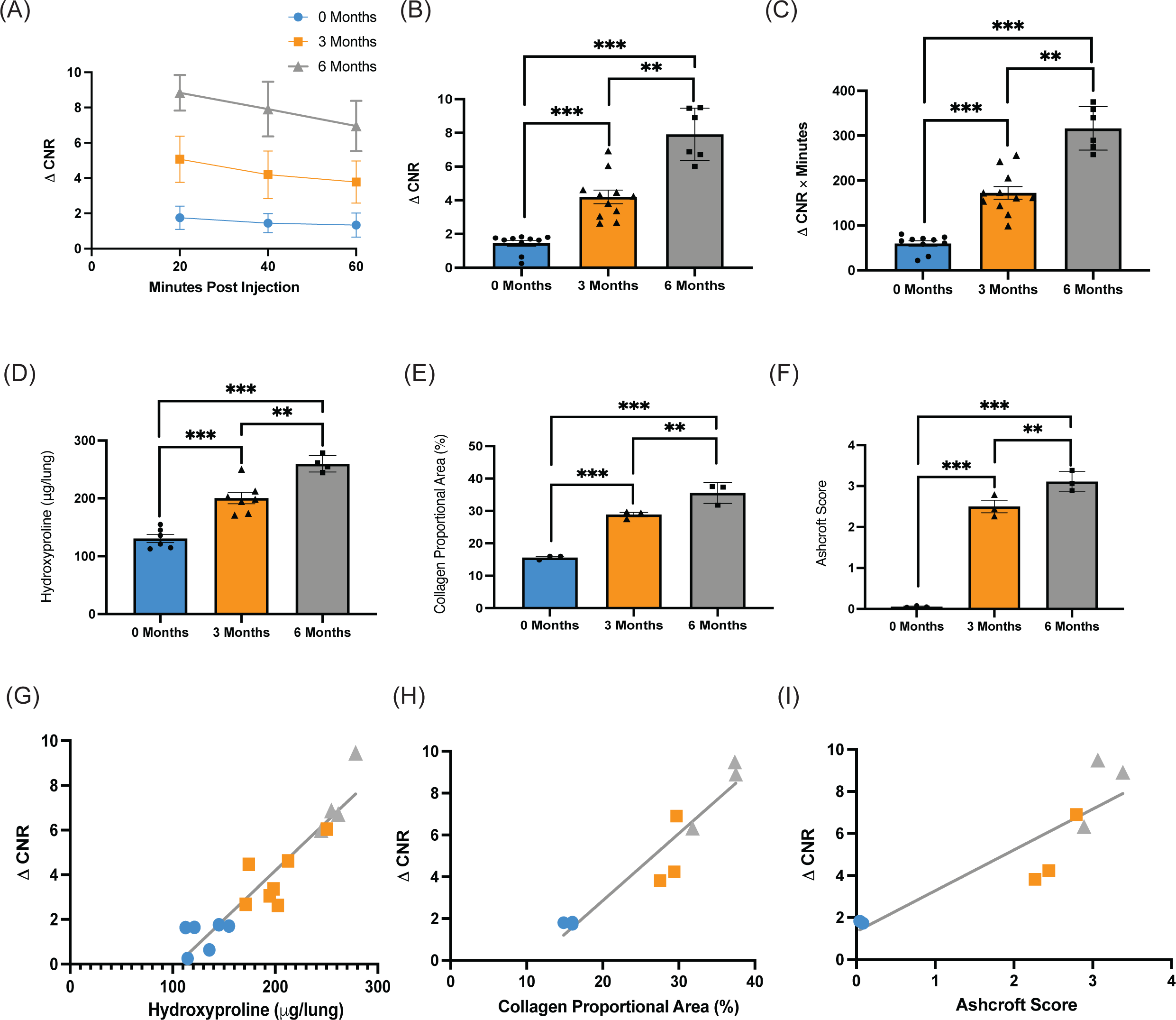
EP-3533 detects progressive RILI in murine model. ΔCNR versus time curves show progressively increased EP-3533 enhanced MR signal at 3 months and 6 months post irradiation (A). ΔCNR at 40 minutes post-injection is progressively elevated from 0 months, 3 months, and 6 months post irradiation (B). Area under the curve quantification also demonstrated similar elevations in EP-3533 signal (C). Validated techniques used to quantify RILI include hydroxyproline (D), collagen proportional area (E), and Ashcroft score (F). Each technique demonstrates increasing severity of RILI over time post irradiation with ANOVA and Bonferroni post hoc t-test. The association between EP-3533 and hydroxyproline (G), collagen proportional area (H), and Ashcroft score (I) were assessed via Pearson correlation and EP-3533 was significantly correlated with RILI severity in each. (* p<0.05, ** p<0.001, *** p<0.0001).

Representative ultrashort echo time (UTE) T1-weighted axial MR images acquired before and 40 minutes after EP-3533 administration, and UTE subtraction images (40 min post-injection – pre-injection) demonstrated increases in right lung signal enhanced by EP-3533 (Figure 1C).

Figure 1D demonstrates co-localization of lung injury in the irradiated right hemithorax. Masson’s trichrome stain shows diffuse injury throughout the right lung but not in the left lung. The T1 weighted coronal MR image shows ground glass opacities in the right hemithorax and normal appearing lung on the left. The subtraction image illustrates substantial signal enhancement in the right lung while little enhancement is observed in the uninjured left lung.

The change in contrast-to-noise ratio (ΔCNR) versus time post EP-3533 injection for 0M, 3M, and 6M post-irradiation groups are shown in Figure 2A. The area under the curve (AUC) for ΔCNR versus time was elevated in the irradiated groups (ANOVA, p<0.001 *post hoc*: 0M 60 ± 6 ΔCNR*min vs 3M 172 ± 14 ΔCNR*min, p<0.001; 0M vs 6M 316 ± 20 ΔCNR*min p<0.001; 3M vs 6M p<0.001) (Figure 2B). The ΔCNR at 40-minutes post-contrast injection has previously been identified as having the largest signal-to-noise ratio in fibrotic tissue^25^, and was elevated following irradiation (ANOVA, p<0.001 *post hoc*: 0M 1.4 ± 0.2 vs 3M 4.2 ± 0.4, p<0.001; 0M vs 6M 7.9 ± 0.6, p<0.001) (Figure 2C).

To test the strength of the association between markers of RILI severity and EP- 3533 signal, disease severity data (CPA, Ashcroft score, hydroxyproline) from each time point was plotted against the corresponding ΔCNR value at 40 minutes post-injection for that animal. Significant correlations were found for each metric (hydroxyproline vs ΔCNR: Pearson r was 0.94, p<0.001) (Figure 2G) (CPA vs ΔCNR: r = 0.95, p<0.001) (Figure 2H), Ashcroft Score vs ΔCNR: r = 0.90, p<0.001) (Figure 2I).

### Effects of losartan on mitigation of RILI

We next sought to test the probe sensitivity to detect disease modification.

Losartan (Los) is an angiotensin receptor blocker with well-known antifibrotic properties, which has been previously shown to improve RILI in both rodent models and in preliminary human studies^16, 18, 37, 38, 39, 40^. Losartan (40 mg/kg/day) was administered in drinking water immediately following irradiation (Figure 3A). At 3 months post-exposure, mice underwent imaging prior to tissue harvest. A visual comparison of the untreated XRT and losartan-treated XRT groups via H&E and Masson’s Trichrome stained sections reveals fewer cellular infiltrates, diminished collagen deposition, and less alveolar thickening in the losartan-treated group (Figure 3B). Representative MR images demonstrate increased probe signal following irradiation, which was diminished with losartan treatment. The unirradiated group and the unirradiated group treated with losartan appeared similar on both imaging and histology.

**Figure 3:**
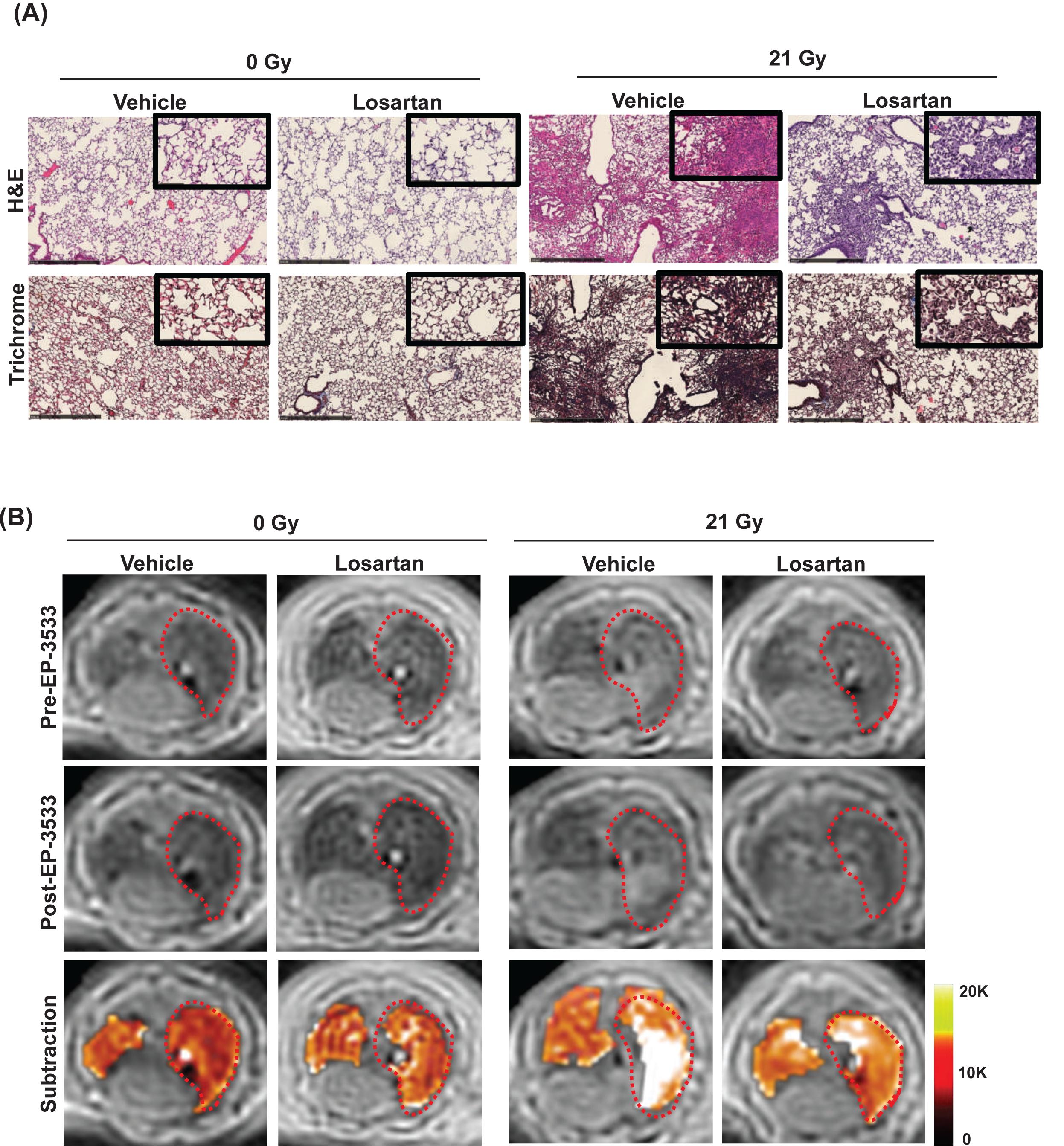
Mitigation of RILI with Losartan in murine model. Representative H&E and trichrome stains of right lung (A) show normal lung architecture in 0 Gy groups while the 21 Gy groups developed interstitial inflammation and alveolar thickening consistent with RILI, which is less severe in losartan treated mice. Representative axial MR images (B) before and after EP-3533 administration. Unirradiated 0Gy mice demonstrate low signal in both lungs, whereas irradiated mice demonstrate increased signal in the right lung consistent with RILI. Images acquired 40 min after EP-3533 administration (B, middle row) show increased lung signal in the irradiated mice compared to the pre-injection images, although signal enhancement is diminished in mice treated with losartan. False color subtraction images (40 minutes following EP-3533 injection – preinjection) overlaid on pre-injection UTE images demonstrate increased signal enhancement in the irradiated lung and the decreased signal enhancement in mice treated with losartan.

Irradiation followed by losartan treatment mitigated RILI but did not eliminate it completely. CPA was elevated in 21 Gy+Los when compared to 0 Gy+Los but was significantly lower than in 21Gy+Veh (ANOVA p<0.001, *post hoc*: 0Gy+Los 17 ± 0.6% vs 21 Gy Los 23 ± 1% p=0.005; 21Gy+Veh 29 ± 0.7% vs 21Gy+Los p=0.009) (Figure 4E). The Ashcroft score was similarly improved with losartan (ANOVA p<0.001, *post hoc*: 0Gy+Los 0.2 ± 0.01 vs 21Gy+Los 1.1 ± 0.06 p=0.018; 21Gy+Veh 2.5 ± 0.15 vs 21Gy+Los p=0.005) (Figure 4F). Hydroxyproline confirmed this trend (ANOVA p<0.001, *post hoc*: 0Gy+Los 143 ± 6 ug/lung vs 21Gy+Los 169 ± 5 ug/lung p=0.022; 21Gy+Veh 201 ± 10 µg/lung vs 21Gy+Los p<0.001) (Figure 4D).

**Figure 4:**
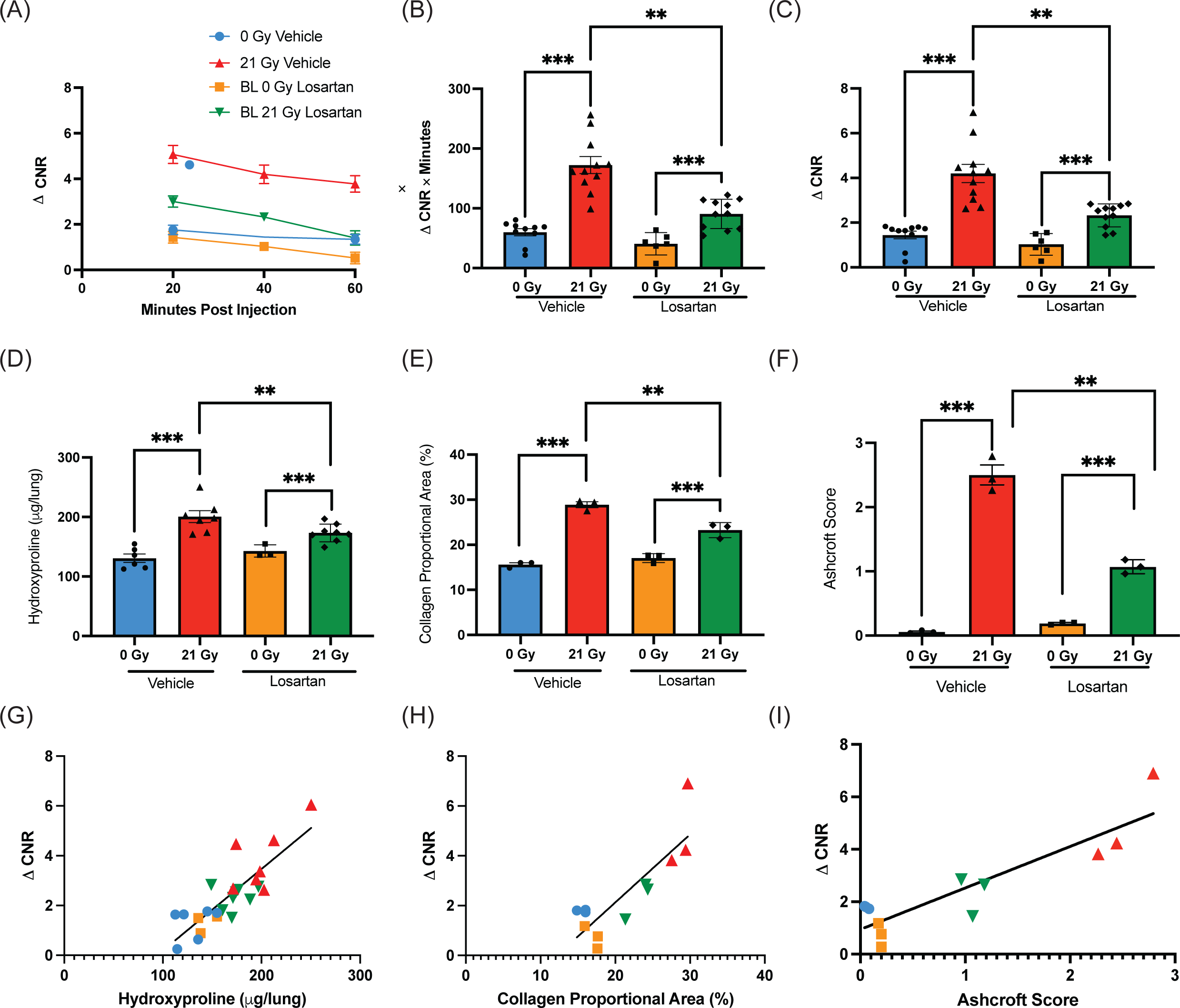
EP-3533 detects pharmacologic mitigation of RILI with losartan in murine model. ΔCNR versus time curves demonstrate increased EP-3533 signal in the irradiated groups, although signal was reduced in the irradiated losartan group (A). Area under the curve quantification (B) and ΔCNR at 40 minutes post-injection (C) revealed elevations in EP-3533 signal in the irradiated vehicle group, which was significantly reduced with losartan treatment. Hydroxyproline, collagen proportional area, and Ashcroft score each show significant increases with radiation, which are reduced in mice treated with losartan. The association between EP-3533 and hydroxyproline (G), collagen proportional area (H), and Ashcroft score (I) were assessed via Pearson correlation and EP-3533 was significantly correlated with RILI severity in each. (* p<0.05, ** p<0.001, *** p<0.0001).

Time-dependent EP-3533 ΔCNR is displayed in Figure 4A. The area under the curve (AUC) analysis (Figure 4B) resolved the increase in signal from irradiation and detected the treatment effect of losartan (ANOVA p<0.001, *post hoc*: 0Gy+Los 41 ± 8 ΔCNR*min vs 21Gy+Los 91 ± 7 ΔCNR*min, p<0.001; 21Gy+Veh 172 ± 14 ΔCNR*min vs 21Gy+Los p<0.001). Similarly, ΔCNR at 40 minutes post-injection confirmed this result (ANOVA p<0.001, *post hoc*: 0Gy+Los 1 ± 0.2 vs 21Gy+Los 2 ± 0.2 p<0.001; 21Gy+Veh 4.2 ± 0.4 vs 21Gy+Los p<0.001) (Figure 4C).

Significant correlations between the MRI metric ΔCNR at 40 minutes post-injection and biochemical and histological measures of fibrosis were found for each metric (Hydroxyproline vs ΔCNR: r = 0.85, p<0.001, Fig 4G; CPA vs ΔCNR: r = 0.84, p<0.001, Fig 4H; Ashcroft Score vs ΔCNR: r = 0.89, p<0.001, Figure 4I). These results confirm a strong correlation between markers of RILI severity and collagen probe signal in the detection of a pharmacological mitigation effect.

### Collagen probe detection of RILI in explanted human tissue

Human lung tissue in subjects with RILI (N=3) and unirradiated subjects (N=3) were harvested and characterized. Tissue donors are described in Supplementary table 1 H&E and Masson’s Trichrome stains show alveolar thickening, cellular infiltration, and collagen deposition in irradiated lung tissue (Figure 5A). Analysis via CPA showed significant increase in irradiated tissue (XRT 39 ± 2 vs Control 28 ± 2 %, P=0.015).

**Figure 5:**
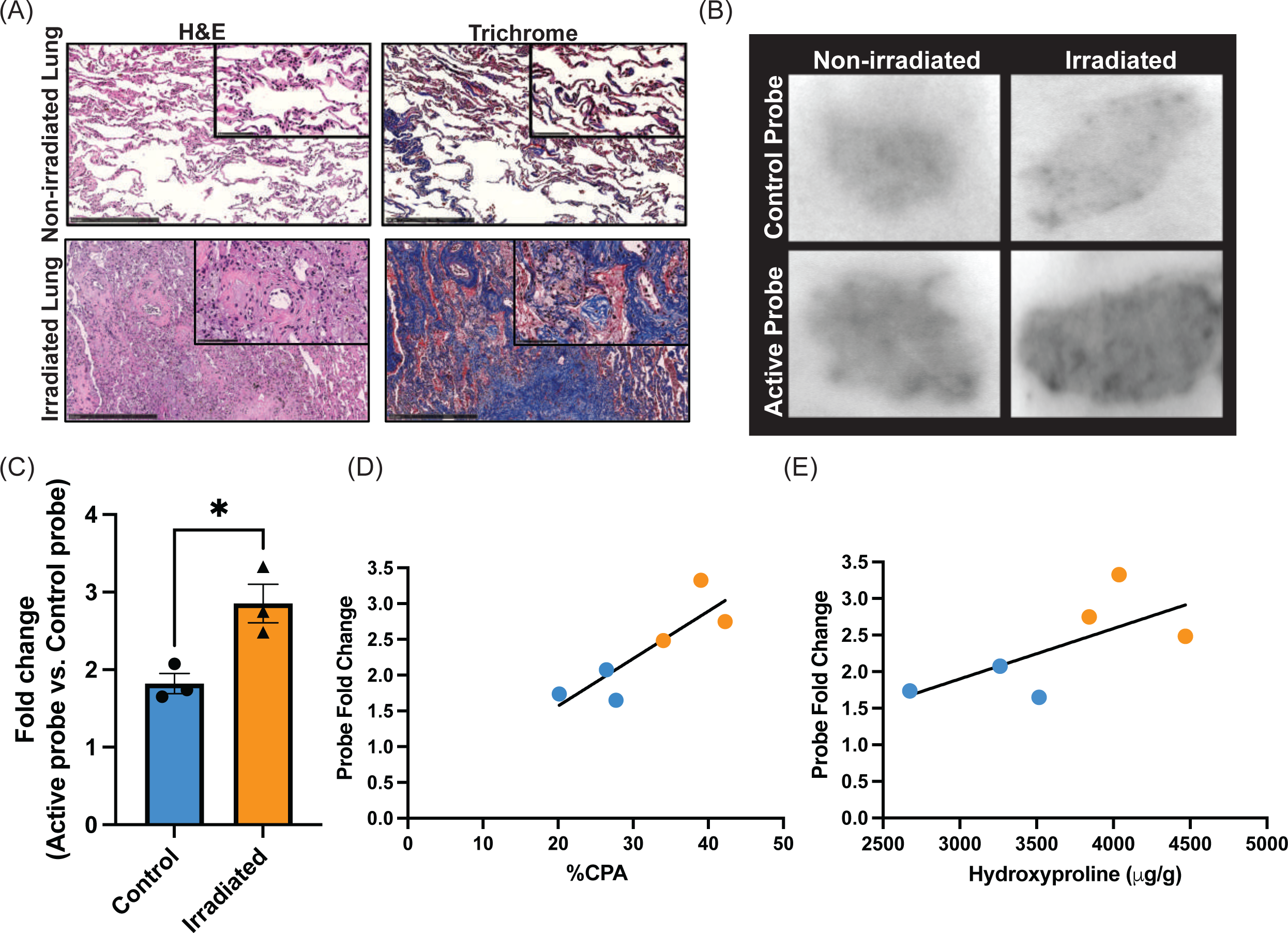
Quantification of 68Ga-CBP8 in human tissue with RILI. H&E and trichrome stains of non-irradiated (top) and irradiated lung (bottom) with characteristic RILI changes including cellular infiltration, alveolar wall thickening, and increased collagen deposition (A). Autoradiography following incubation with 68Ga-ctrl shows similar uptake between irradiated and non-irradiated lung tissue (upper panels). 68Ga-CBP8 has increased uptake in irradiated vs non-irradiated lung tissue (lower panels) (B). 68Ga- CBP8 intensity normalized to 68Ga-ctrl signal intensity is significantly increased in irradiated tissue p=0.021 (C) and is significantly correlated with CPA p=0.026 (D), but not with hydroxyproline (E).

Hydroxyproline was also significantly elevated in irradiated tissue (Control 3150 ± 249 vs XRT 4116 ± 185 ug/g P=0.036). Signal from human lung tissue sections incubated with ^68^Ga-CBP8 was significantly increased in RILI tissue (XRT 11807 ± 789 vs control 8584 ± 95 DLU P=0.015). The ^68^Ga-CBP8 signal was normalized to the signal from a nonbinding linear peptide control probe, ^68^Ga-Ctrl, to further assess specificity. The ratio of ^68^Ga-CBP8 to ^68^Ga-Ctrl autoradiography signal was significantly higher in the RILI tissue (Control 1.8 ± 0.1 vs XRT 2.9 ± 0.3 vs P= 0.021 Fig 5C). Representative images of probe uptake are shown in Figure 5B.

The strength of the relationship between ^68^Ga-CBP8 and each RILI severity metric was assessed. A significant correlation was found for CPA (CPA vs fold change: r = 0.87, p=0.026, Figure 5D) but hydroxyproline was not significantly associated with fold change (hydroxyproline vs fold change: r = 0.67, p value >0.05, Figure 5E).

### In vivo imaging of RILI in humans with ^68^Ga-CBP8

We obtained permission to evaluate RILI in subjects who underwent radiation therapy as part of routine treatment for lung cancer. Six subjects were recruited and imaged successfully. Mean age 73 (range 64-79), 17% male, BMI 26 ± 2.2, tumor type: 44% squamous cell carcinoma, 66% adenocarcinoma, pack-years 39 ±11, Common Terminology Criteria of Adverse Events v5 1.7 ± 0.3 (Table 1). Figure 6 presents representative images of three subjects. CT images of the irradiated lung confirm RILI. MR images also demonstrate lung injury. Fused PET-MR images demonstrate increased ^68^Ga-CBP8 uptake in the area of RILI. Subject 1 received ^68^Ga-CBP8 while in the scanner, and the probe’s dynamic washout curve was measured (Supplementary Figure 2).

**Figure 6:**
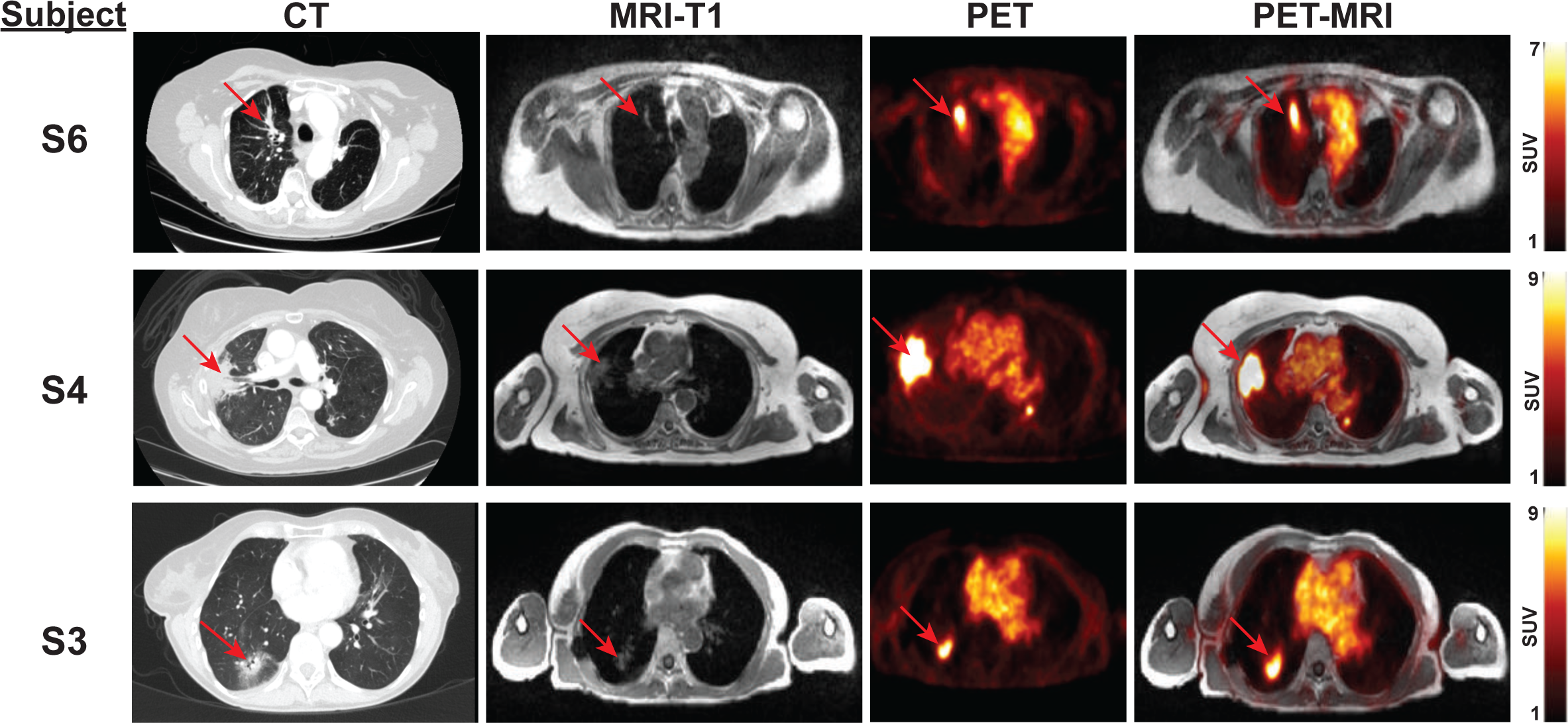
Representative matching axial CT, MR, PET, and PET-MRI images of human subjects with RILI. Each row represents an individual subject. CT images (left) depict RILI in irradiated lung with characteristic consolidation and ground glass opacities. Red arrows indicate areas of RILI. Corresponding T1-weighted MR images obtained simultaneously with the PET study (second column). PET (third column) and fused PET-MRI images (fourth column) demonstrate elevated signal in areas of RILI following the injection of 68Ga-CBP8.

**Table 1:**
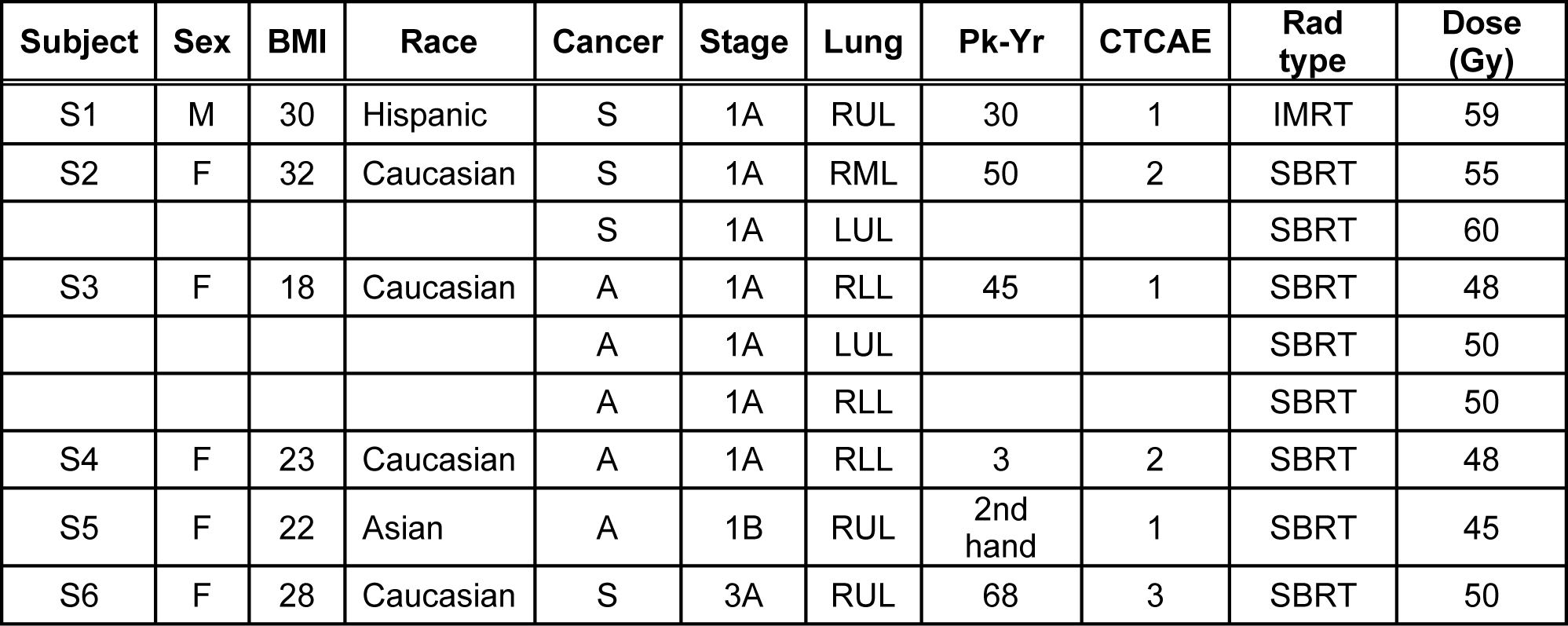
Human subjects who underwent imaging with type 1 collagen PET probe. Six subjects were imaged. S2 and S3 had undergone irradiation to multiple tumors and each lesion was analyzed separately to give nine RILI lesions. (S) Squamous cell carcinoma, (A) Adenocarcinoma, (Pk-Yr) smoking pack-years, (CTCAE) Common Terminology Criteria of Adverse Events v5, (SBRT) Stereotactic Body Radiation Therapy, (IMRT) Intensity Modulated Radiation Therapy. No subjects received concurrent chemotherapy.

The area of RILI was identified and agreed upon by two independent radiologists reviewing the images. The mean standardized uptake values (SUV) 40-75 minutes post-injection were computed and compared with areas of contralateral healthy lung.

Mean SUV in areas of RILI (1.23 ± 0.19) was significantly increased over that of healthy lung (0.35 ± 0.04, p<0.001).

The 35-minute PET acquisition was binned into 5-minute frames. SUV_mean_ for RILI, healthy lung, and blood pool ROIs were quantified to produce washout curves (Figure 7A). The tissue-to-blood ratio was calculated for both RILI and healthy lung ROIs graphed against time after injection (Figure 7B). This graph produces a flat curve and suggests that the probe is stable at this point in its washout curve, and appropriate for analyzing the probe signal. The mean SUV_mean_ signal from 40-75 minutes post-injection for the RILI ROI was significantly higher than the healthy lung ROI (1.2 ± 0.2 vs 0.4 ± 0.1 p=0.005), (Figure 7C). The tissue-to-blood ratio for RILI was also significantly increased over healthy lung ROI (0.64±0.06 vs 0.19±0.02 p<0.0001) (Figure 7D). These results indicate a greater affinity of ^68^Ga-CBP8 for areas of RILI over healthy lung.

**Figure 7:**
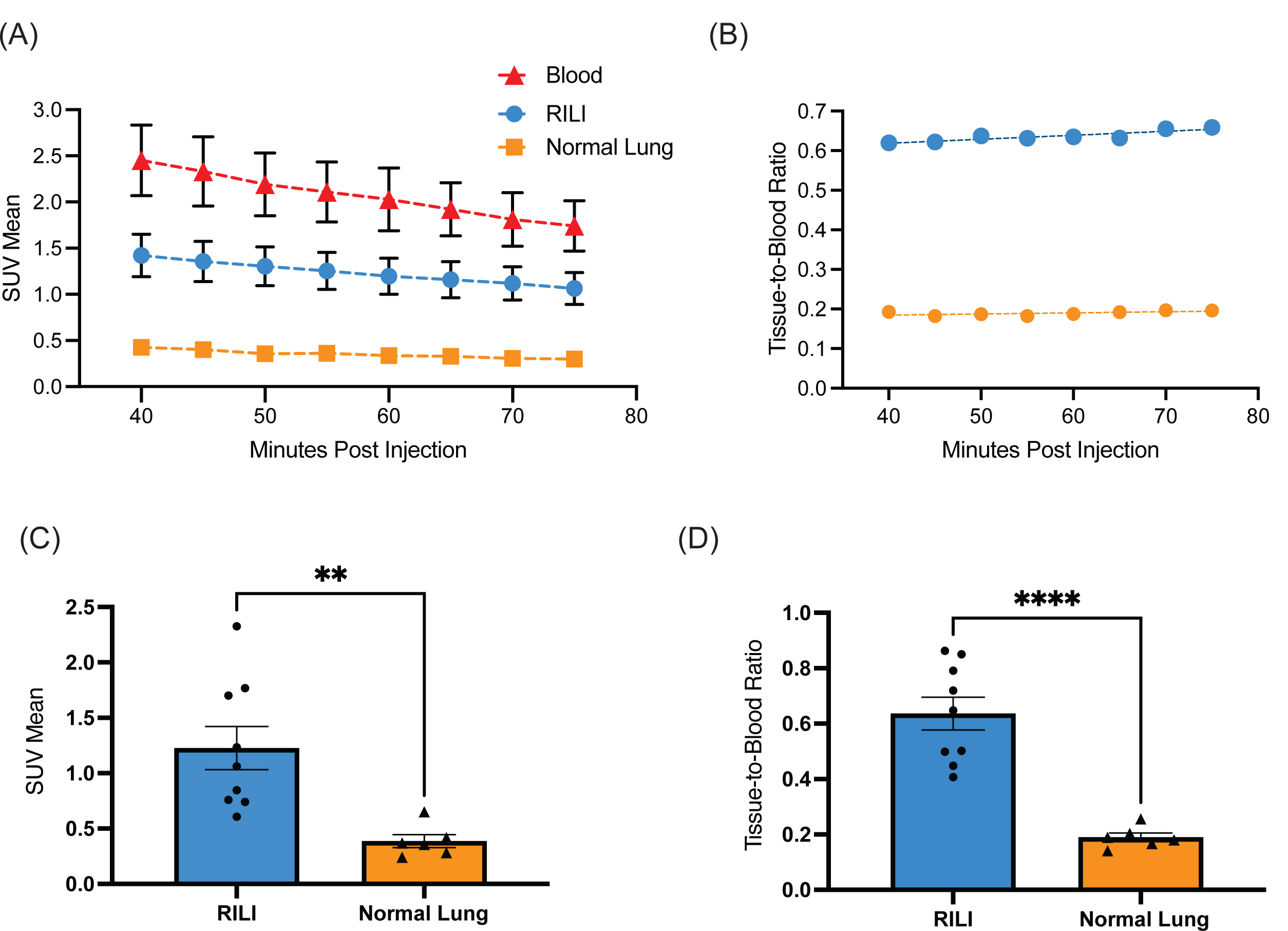
Quantitative analysis of 68Ga-CBP8 PET images in human subjects with RILI. (A) 68Ga-CBP8 washout curves derived from dynamic PET images in areas of RILI, healthy lung, and blood pool. (B) Tissue-to-blood ratio for RILI and healthy lung. (C) Mean SUV is significantly elevated in RILI vs normal lung, p=0.005. (D) Tissue-to-blood ratio is significantly elevated in RILI vs normal lung, p<0.001.

## Discussion

We demonstrated in a pre-clinical model of RILI that noninvasive molecular imaging techniques utilizing a probe specific to type 1 collagen, a building block of fibrotic tissue in the lung, is sensitive to changes in RILI – both with the natural progression of disease over time and with pharmacological disease modification. Type 1 collagen molecular MR imaging with EP-3533 was highly correlated with established markers of RILI such as hydroxyproline and histological analysis. Because this MR probe is not available for human use, we next used an analagous PET probe, ^68^Ga- CBP8, that employed the same collagen-targeting peptide and that was available for human studies. We tested the binding and specificity of ^68^Ga-CBP8 to RILI in human tissue using autoradiography and showed that the collagen probe signal was again highly correlated with RILI severity. Finally, we demonstrated the ability of ^68^Ga-CBP8 to detect RILI in human subjects *in vivo*. This approach to molecular detection of collagen deposition in irradiated lungs is novel and has not been described in humans.

Murine models of RILI are well established in the literature^36^. RILI in mice closely mimics disease in humans. It is slowly progressive over time with early pneumonitis and chronic fibrosis stages^41^. RILI appears to share common signaling pathways with other fibrotic lung diseases^42^. RILI models take months to develop and are frustratingly slow in comparison to bleomycin models. However, because they are slow moving and mimic human disease, RILI models may be ideal for the study of novel antifibrotic therapeutics which may be effective in many forms of fibrotic lung disease. Murine models of pulmonary fibrosis and fibrogenesis are a commonly used preclinical model for the development of molecular imaging probes^43^. Our group has previously tested probes targeted to type 1 collagen in a bleomycin model, as well as another probe directed at a key step in collagen synthesis^27, 34, 44, 45^. However, this is the first attempt to explore molecular imaging techniques in preclinical RILI models.

Inhibition of the renin-angiotensin pathway has been studied as a mitigation agent in preclinical models of RILI^18, 37^ and has been associated with reduced rates of radiation induced pneumonitis in humans. In two small retrospective studies angiotensin-converting enzyme inhibitor (ACEI) use was associated with a significant reduction in symptomatic radiation-induced pneumonitis from a baseline of 16% to 4% in on study and from 11% to 2% in the other^16, 38^. Another retrospective study showed less symptomatic radiation-induced pneumonitis in subgroup analysis of males and low dose radiation^40^. A prospective trial was attempted but closed early due to low subject accrual^39^. These studies illustrate the need for more sophisticated methods to evaluate RILI.

In humans, molecular imaging techniques have been successfully applied to idiopathic pulmonary fibrosis, with molecular probes directed at various aspects of fibrosis including: type 1 collagen^28^, chemokine receptor-2^46^, αVβ6 integrin^47^, cathepsin protease^48^, and glycolytic metabolism^49^. However, molecular imaging studies have yet to focus on RILI. Significant work has been done to quantify RILI using more traditional techniques^50^. The study of RILI via noninvasive molecular imaging techniques represents a unique opportunity, as the timing, location, and dose are carefully recorded, and the patients are carefully followed thereafter. Noninvasive quantification of RILI offers the possibility of utilizing molecules specific to the disease process, such as type 1 collagen to resolve subtle changes in disease with therapeutics that are undetectable with current methods. Another potential application is the identification of at-risk individuals who will go on to develop severe disease and may benefit from preemptive therapy and close follow-up.

^68^Ga-CBP8 is well tolerated in human subjects to date. It binds to its target quickly and is rapidly cleared from the systemic circulation and healthy tissue. Ga-68 has a half-life of 68 minutes, minimizing radiation exposure. In animal studies and in humans it has been demonstrated that ^68^Ga-CBP8 binds to disorganized collagen associated with fibrosis but does not accumulate in organized, normal collagen such as bone.

This targeted molecular imaging probe, ^68^Ga-CBP8, can detect subtle changes in RILI progression (potentially improving upon routine CT chest) and provide important prognostic information for the development of RILI and other fibrotic diseases. While not yet tested in humans, this probe is capable of detecting treatment-induced attenuation of RILI in mice. This technique could also provide a highly sensitive assay for therapeutic drug trials – an unmet need that will be essential to improve the efficacy of novel therapeutics for RILI. By resolving subtle changes in early disease, this molecular imaging strategy may identify subjects at high risk for severe RILI before they develop symptoms. Finally, this technology is being evaluated to distinguish post-radiation change from cancer recurrence in a murine model of lung cancer treated with radiation. In mice, this technology offers the exciting possibility of noninvasively assessing disease burden at multiple time points throughout an experiment.

The findings of this study need to be interpreted in light of its small sample size.

Additional studies are needed to characterize these preliminary findings more thoroughly. More work is required to fully correlate ^68^Ga-CBP8 signal with specific histopathological findings, and to develop the association between hydroxyproline and ^68^Ga-CBP8 into a robust quantitative method. We intend to image subjects undergoing pneumonectomy to correlate imaging findings with disease in the explanted lung.

Subjects were imaged utilizing MR/PET – which makes comparison of our findings with clinical CT scans via co-registration difficult. Ideally the ^68^Ga-CBP8 probe needs to be evaluated with a co-registered CT-PET. As these findings represent a snapshot in time, further studies and repeat imaging in the same subject are needed to correlate probe signal with disease progression.

We conclude that the noninvasive monitoring of type 1 collagen via molecular imaging techniques in both clinical and preclinical RILI will enhance our assessment of the disease. This technique offers new opportunities to noninvasively monitor RILI, with potential applications in both disease prognosis and in assessment of treatment response. This study is the first step in expanding techniques that have been focused on traditional pulmonary fibrosis into the study of RILI.

## Methods

### Murine RILI Model

All experiments and procedures were performed in accordance with the National Institutes of Health’s “Guide for the Care and Use of Laboratory Animals” and were approved by the Massachusetts General Hospital Institutional Animal Care and Use Committee (protocol # 2010N000005). C57L/J (Stock # 000668) male mice aged 12 weeks (Jackson Laboratories, Barr Harbor, ME) were obtained. A total of 44 mice were used in this study.

### Irradiation

Irradiated mice were anesthetized with 87.5 mg/kg ketamine and 12.5 mg/kg xylazine via intraperitoneal injection and secured in custom lead shielding prior to right hemithorax irradiation of 21 Gy via dual source ^137^Cs Gammacell 40 Exactor (Best Theratronics, Ottawa, Ontario). Following irradiation, animals were returned to their cages and allowed to recover. Mice were kept in a clean animal facility with ad libitum access to food (Purina Test Diet, Richmond, IN USA) and acidified water with 12-hour light/dark cycle. In the losartan treatment groups, mice were allowed ad libitum access to losartan (American Health Packaging, Columbus, Ohio) 40 mg/kg/day dissolved in standard acidified water (vehicle) immediately following irradiation^30, 31^. Water bottles were monitored and refreshed frequently.

### Radiation Shielding Validation

Thermoluminescent dosimeters (TLDs) were used to measure the dose delivered via dual source ^137^Cs Gammacell 40 Exactor (Best Theratronics, Ottawa, Ontario) to the center of an opening in a custom-made lead shielding apparatus. A series of paired dosimeters were irradiated in the center of the empty specimen cannister of the Cesium irradiator (Supplementary Table 2). Four pairs of TLDs were irradiated over a range of doses from 0.5 Gy to 2.0 Gy, as set by a timer setting on the irradiator calculated from the machine’s daily dose rate which is derived from the initial activity of the Cs source and its decay rate. This was done to measure the output of the irradiator and create a dose response curve to characterize how the TLDs record dose in the Cs irradiator in an open configuration. Next a series of paired dosimeters were irradiated in the opening of the lead shielding apparatus. They were enclosed in the center of 1.2 cm thick tissue equivalent material to simulate midplane in a mouse body. This was done for an open configuration dose of 1.0 Gy and 2.0 Gy as prescribed by the timer setting given by the machine’s daily dose rate. The ratio of the measured open configuration dose to that of the in-shield dose for a given machine time setting was used to calculate the dose that the mouse lung received, which was 76% of the open field dose (Supplementary Figure 1). From this we infer that a 21 Gy exposure from the irradiator resulted in a 16 Gy tissue dose to the right lung through the aperture of the shielding.

### MR Imaging

Imaging protocol as previously described^22^. Briefly, prior to imaging animals were anesthetized with 1–2% isoflurane in 30% oxygen and secured in custom cradle. The tail vein was cannulated and used for injection of probe. Respiration rate was monitored by a small animal physiological monitoring system (SA Instruments Inc., Stony Brook NY) with target 60 breaths/min. Imaging was performed using a small-bore 4.7 T animal scanner (Biospec, Bruker, Billerica MA) with a custom-built volume coil. T1-weighted (T1w) 3D Ultra Short Time to Echo (UTE) images (repetition time (TR) / echo time (TE) = 4/0.008 ms; flip angle, 5°; field of view, 75 x 75 x 75 mm3; matrix size: 128 x 128 x 128; spatial resolution = 586 μm isotropic; acquisition time, 3 minutes 23 seconds) were obtained prior to and at 20-, 40-, and 60-minutes after the injection of 3 µmol/kg EP- 3533. During and immediately following probe injection, a series of 10 T1w 3D Fast Low Angle Shot (FLASH): images (TR/TE = 20/3 ms; flip angle, 12°; field of view, 85 x 65 x 50 mm3; matrix size: 212 x 162 x 100; spatial resolution = 400 x 400 x 500 μm3; acquisition time, 3 minutes 17 seconds) were acquired to monitor probe injection and clearance.

### Molecular Probes

EP-3533 comprises a 10–amino acid, disulfide bridged cyclic peptide conjugated to three Gd moieties with affinity for type I collagen (Kd = 1.8 µM) and strong MR signal enhancement (relativity, r1 = 16.2 mM^-1^s^-1^ [5.4/Gd ion] at 4.7T), which was synthesized as described previously^20^. ^68^Ga-CBP8 utilizes the same peptide as EP-3533 but is functionalized with a NODAGA chelator for labeling with Ga-68 A non-binding control probe, ^68^Ga-Ctrl, was prepared by reduction of the peptide disulfide and alkylation of the thiol groups with iodoacetamide, similarly to a previously described procedure^32^; the resultant linear peptide has negligible affinity for type I collagen^21^. The specificity of EP- 3533 for type 1 collagen has been demonstrated *in vivo* previously^22^.

### Murine MR Image Analysis

Images were analyzed using the program Horos (www.horosproject.org). Regions of Interest (ROI) were identified on the lung, back muscle, and air adjacent to the animal of 5 contiguous slices of pre and post contrast UTE images. Signal intensity (SI) of the lung and muscle of each slice was obtained and standard deviation (SD) of the signal in the air adjacent to the animal was used to estimate noise. For each slice, contrast to noise ratio (CNR) was calculated as follows: (SI_lung_ – SI_muscle_)/SD_air_. An average CNR of all image slices was calculated for the pre-injection images (CNR_pre_) and for the post-injection images (CNR_post_) at 20-, 40-, and 60-minutes post injection. The change in CNR, ΔCNR = CNR_post_ – CNR_pre_, represents lung enhancement due to collagen targeting.

### Tissue Analysis

Following imaging, animals were euthanized, and the lungs were harvested. For each experimental condition, a subset of harvested lungs was frozen for molecular analysis. Lung tissue was homogenized and hydroxyproline was quantified by HPLC as previously described^33^. Another subset underwent inflation and fixation of the lungs in neutral 10% buffered formalin, embedded in paraffin, and sectioned into 5 mm-thick slices for hematoxylin and eosin (H&E), Masson’s Trichrome, and Sirius red with fast green staining.

Collagen Proportional Area (CPA) and Ashcroft Score:

Sirius red stained slides were digitally scanned and images were analyzed by thresholding to determine the collagen proportional area (CPA), % area stained collagen positive, using ImageJ (National Institute of Health, Bethesda MD) as previously described^34^. Masson’s Trichrome stained sections were analyzed by a pathologist (LH), who was blinded to the study, to score the amount of lung disease via the Ashcroft method^35^.

### Human Tissue and Autoradiography

With Partners Institutional Review Board approval (# 2014P000559), remnant human lung resection specimens were obtained from subjects who received radiation therapy for lung cancer (N=3) and nonirradiated control subjects (N=3) (Supplementary Table 1). Tissue was then fixed in OCT and frozen sections 10 µm thick were obtained.

Sections were thawed and washed in PBS for 5 minutes three times. The active probe against type 1 collagen, ^68^Ga-CPB8, and the inactive control probe ^68^Ga-Ctrl were radiolabeled as described previously^27^. Radioactivity of both probes was quantified using a dose calibrator (Capintec, CRC-55tR, NJ) and found to be similar (1.54 mCi). Tissue sections were incubated with equal radiochemical concentrations of either ^68^Ga- CBP8 or nonbinding ^68^Ga-Ctrl for 45 minutes in PBS. The sections were then washed three times with PBS for 5 min. The residual tissue radioactivity was then quantified via Cyclone Plus Phosphor Storage system (Perkin Elmer, Waltham, MA). Analysis performed with Image-J software and quantified with mean arbitrary units.

### *In Vivo* Human Imaging

This study was approved by the Partners Institutional Review Board IRB # 2020P001899, #2017P002718 and registered at clinicaltrials.gov (identifier: NCT04485286, NCT03535545).

Inclusion criteria: Age 18-80, stage I – III NSCLC, received radiation therapy as routine care for lung cancer, diagnosed with radiographically apparent RILI by treating physician.

Exclusion criteria: Subjects were ineligible to participate if: eGFR <30, Stage 4 cancer, pregnant or breastfeeding, BMI >33, known history of pulmonary disease, active smoker, pulmonary infection in past 6 weeks, contraindication to MRI.

### PET-MR Imaging Protocol

Subjects provided informed consent. Following a physical exam, a peripheral IV was placed and ^68^Ga-CBP8 was administered. Approximately 173 MBq (range 148–236 MBq) of ^68^Ga-CBP8 was administered. Subject 1 received ^68^Ga-CBP8 while position in the PET-MRI (3 Tesla Biograph mMR, Siemens Healthineers, Erlangen, Germany) to evaluate probe kinetics. All other subjects were placed in the MRI 30 minutes post injection. Simultaneous ^68^Ga-CBP8 PET and MRI of the thorax was performed.

Emission data were acquired for up to 90 minutes in listmode format after the injection of ^68^Ga-CBP8. Attenuation correction was performed using the manufacturer’s MR-based method. Images corresponding to 5-minute dynamic frames were reconstructed using the standard reconstruction algorithm (OSEM, 21 subsets, three iterations, 256 × 256 matrix size,127 slices) and smoothed with a 4-mm full width at half maximum Gaussian filter. Simultaneous to the PET data acquisition the MR data were acquired with several sequences, including T2-weighted STIR HASTE images acquired coronally during 2 concatenated breath-holds using the following parameters: TE/TR = 100/1000 ms; TI = 0.2 s; flip angle = 120°; reconstruction matrix = 256x179x36, 1.56x1.56x6.9 mm^3^; time of acquisition = 36 s. Additionally, dual-echo Dixon images were acquired in order to enable attenuation correction of the PET images, as standard on the mMR scanner, with the following parameters: TE = 1.23/2.46 ms; TR = 3.6 ms; flip angle = 10°; reconstruction matrix = 192x126x128, 2.6x2.6x2.23 mm^3^; acquisition time = 19 s.

Subjects were contacted the following day to assess for any adverse effects – none reported.

### Image analysis

Two independent radiologists reviewed the most recent clinical CT scans of each subject and identified a ROI corresponding to the RILI, and a contralateral ROI of similar size was identified with normal appearing lung. PET signal from 40 to 75 minutes post probe injection was quantified in the ROI’s containing both RILI and normal lung. PET signal was binned in 5-minute intervals and quantified in the ROI’s containing both RILI and normal lung to provide a washout curve. Blood pool PET signal was quantified in the left ventricle.

### Statistics

Statistical analysis was performed using GraphPad (Prism version 6.0). Intergroup analysis performed with one-way ANOVA with post hoc t-test and Bonferroni correction. Area under the curve analysis was performed using the trapezoid method. Strength of linear relationship measured via Pearson correlation. RILI ROI comparison with contralateral healthy tissue was performed with a paired t-test. *P* value < 0.05 considered significant. Data are reported as mean ± SE.

## Data Availability

All data produced in the present work are contained in the manuscript

## Acknowledgements

We would like to thank Dr. John Moulder of the Medical College of Wisconsin for his advice regarding radiation injury models and Dr. Alexsia Richards of the Whitehead institute for her assistance with figures.

## Author contributions

All authors provided critical review of the work, all approved the final version submitted, and all agree to be accountable for the work. Additionally:

Eric Abston: Project conceptualization, data analysis, manuscript preparation

Iris Zhou: Data analysis

Jonathan Saenger: Data analysis

Sergey Shuvaev: Radiolabeling, chemistry

Eman Akam: molecular probe chemistry

Shadi Esfahani: data analysis, figure preparation

Lida Hariri: data analysis, pathology subject matter expert

Nicholas Rotile: data acquisition

Elizabeth Crowley: data acquisition

Sydney Montesi: clinical trial support

Valerie Humblet: project conceptualization

Melin Khandekar: project conceptualization

Grae Arabasz: clinical trial technical support

Ciprian Catana: technical support

Florian Fintelmann: data analysis

Peter Caravan: project conceptualization

Michael Lanuti: project conceptualization, data analysis, funding acquisition, manuscript review

## Data and materials availability

All data are available in the main text or the supplementary materials.

## Online Supplement

### Supplementary Methods

#### Murine RILI Model

All experiments and procedures were performed in accordance with the Declaration of Helsinki and the National Institutes of Health’s “Guide for the Care and Use of Laboratory Animals” and were approved by the Institutional Animal Care and Use Committee. C57L/J (Stock # 000668) male mice aged 12 weeks (Jackson Laboratories, Barr Harbor, ME) were obtained. A total of 44 mice were used in this study.

#### Irradiation

Irradiated mice were anesthetized with 87.5 mg/kg ketamine and 12.5 mg/kg xylazine via intraperitoneal injection and secured in custom lead shielding prior to right hemithorax irradiation of 21 Gy via dual source ^137^Cs Gammacell 40 Exactor (Best Theratronics, Ottawa, Ontario). Following irradiation, animals were returned to their cages and allowed to recover. Mice were kept in a clean animal facility with ad libitum access to food (Purina Test Diet, Richmond, IN USA) and acidified water with 12-hour light/dark cycle. In the losartan treatment groups, mice were allowed ad libitum access to losartan (American Health Packaging, Columbus, Ohio) 40 mg/kg/day dissolved in standard acidified water (vehicle) immediately following irradiation^1, 2^. Water bottles were monitored and refreshed frequently.

#### MR Imaging

Imaging protocol as previously described^3^. Briefly, prior to imaging animals were anesthetized with 1–2% isoflurane in 30% oxygen and secured in custom cradle. The tail vein was cannulated and used for injection of probe. Respiration rate was monitored by a small animal physiological monitoring system (SA Instruments Inc., Stony Brook NY) with target 60 breaths/min. Imaging was performed using a small-bore 4.7 T animal scanner (Biospec, Bruker, Billerica MA) with a custom-built volume coil. T1-weighted (T1w) 3D Ultra Short Time to Echo (UTE) images (repetition time (TR) / echo time (TE) = 4/0.008 ms; flip angle, 5°; field of view, 75 x 75 x 75 mm3; matrix size: 128 x 128 x 128; spatial resolution = 586 μm isotropic; acquisition time, 3 minutes 23 seconds) were obtained prior to and at 20-, 40-, and 60-minutes after the injection of 3 µmol/kg EP- 3533. During and immediately following probe injection, a series of 10 T1w 3D Fast Low Angle Shot (FLASH): images (TR/TE = 20/3 ms; flip angle, 12°; field of view, 85 x 65 x 50 mm3; matrix size: 212 x 162 x 100; spatial resolution = 400 x 400 x 500 μm3; acquisition time, 3 minutes 17 seconds) were acquired to monitor probe injection and clearance.

#### Molecular Probes

EP-3533 comprises a 10–amino acid, disulfide bridged cyclic peptide conjugated to three Gd moieties with affinity for type I collagen (Kd = 1.8 µM) and strong MR signal enhancement (relativity, r_1_ = 16.2 mM^-1^s^-1^ [5.4/Gd ion] at 4.7T), which was synthesized as described previously^4^. ^68^Ga-CBP8 utilizes the same peptide as EP-3533 but is functionalized with a NODAGA chelator for labeling with Ga-68 A non-binding control probe, ^68^Ga-Ctrl, was prepared by reduction of the peptide disulfide and alkylation of the thiol groups with iodoacetamide, similarly to a previously described procedure^5^; the resultant linear peptide has negligible affinity for type I collagen^6^. The specificity of EP- 3533 for type 1 collagen has been demonstrated *in vivo* previously^3^.

#### Tissue Analysis

Following imaging, animals were euthanized, and the lungs were harvested. For each experimental condition, a subset of harvested lungs was frozen for molecular analysis. Lung tissue was homogenized and hydroxyproline was quantified by HPLC as previously described^7^. Another subset underwent inflation and fixation of the lungs in neutral 10% buffered formalin, embedded in paraffin, and sectioned into 5 mm-thick slices for hematoxylin and eosin (H&E), Masson’s Trichrome, and Sirius red with fast green staining.

#### Collagen Proportional Area (CPA) and Ashcroft Score

Sirius red stained slides were digitally scanned and images were analyzed by thresholding to determine the collagen proportional area (CPA), % area stained collagen positive, using ImageJ (National Institute of Health, Bethesda MD) as previously described^8^. Masson’s Trichrome stained sections were analyzed by a pathologist (LH), who was blinded to the study, to score the amount of lung disease via the Ashcroft method^9^.

#### Human Tissue and Autoradiography

With Institutional Review Board approval, remnant human lung resection specimens were obtained from subjects who received radiation therapy for lung cancer (N=3) and nonirradiated control subjects (N=3) (Supplementary Table 1). Tissue was then fixed in OCT and frozen sections 10 µm thick were obtained. Sections were thawed and washed in PBS for 5 minutes three times. The active probe against type 1 collagen, ^68^Ga-CPB8, and the inactive control probe ^68^Ga-Ctrl were radiolabeled as described previously^10^. Radioactivity of both probes was quantified using a dose calibrator (Capintec, CRC-55tR, NJ) and found to be similar (1.54 mCi). Tissue sections were incubated with equal radiochemical concentrations of either ^68^Ga-CBP8 or nonbinding ^68^Ga-Ctrl for 45 minutes in PBS. The sections were then washed three times with PBS for 5 min. The residual tissue radioactivity was then quantified via Cyclone Plus Phosphor Storage system (Perkin Elmer, Waltham, MA). Analysis performed with Image-J software and quantified with mean arbitrary units.

#### *In Vivo* Human Imaging

This study was approved by the Institutional Review Board and registered at <u>clinicaltrials.gov</u>.

#### PET-MR Imaging Protocol

Subjects were contacted the following day to assess for any adverse effects – none reported.

## Supplementary Results

**Supplementary Table 1:**
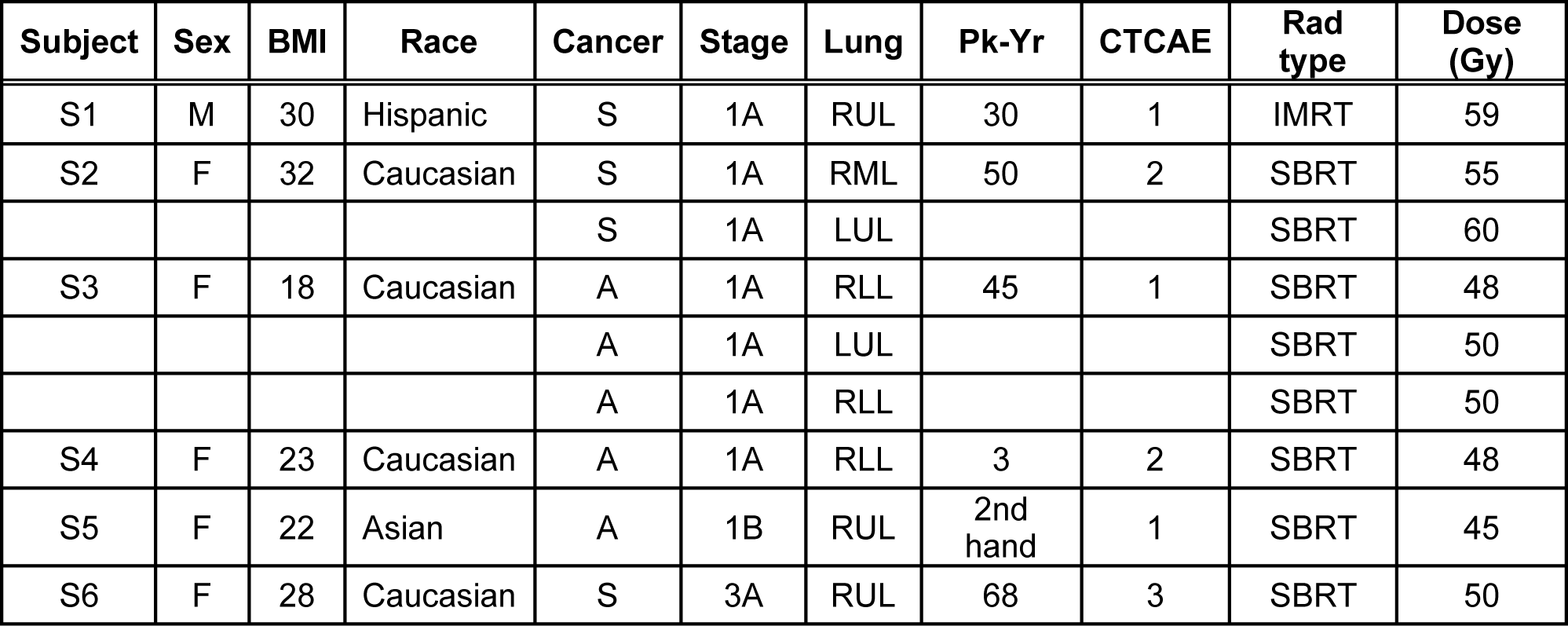
Human subjects who underwent imaging with type 1 collagen PET probe. Six subjects were imaged. S2 and S3 had undergone irradiation to multiple tumors and each lesion was analyzed separately to give nine RILI lesions. (S) Squamous cell carcinoma, (A) Adenocarcinoma, (Pk-Yr) smoking pack-years, (CTCAE) Common Terminology Criteria of Adverse Events v5, (SBRT) Stereotactic Body Radiation Therapy, (IMRT) Intensity Modulated Radiation Therapy. No subjects received concurrent chemotherapy.

**Supplementary Table 2:**
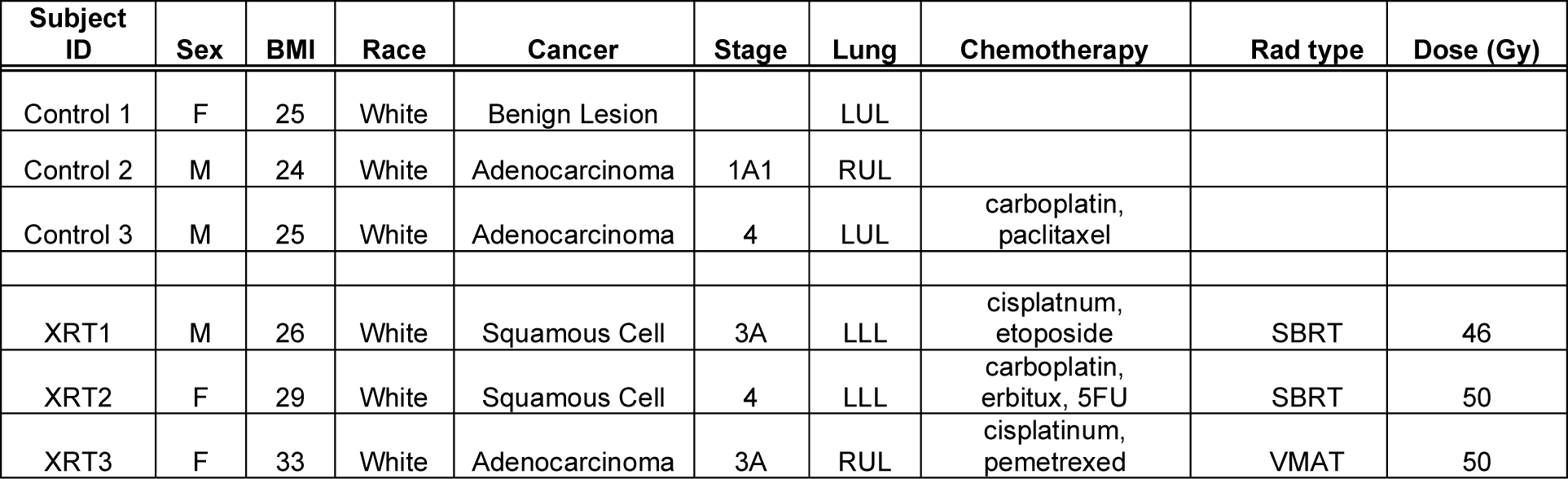
Characteristics of human tissue donors of lung tissue used in autoradiography experiments described in Fig 5. “Lung” column describes region of lung where tissue originated from. “Chemotherapy” column reports chemotherapeutic regimens administered prior to biopsy. (SBRT) Stereotactic Body Radiation Therapy, (VMAT) Volumetric Modulated Arc Therapy. Unirradiated “control” subjects (N=3)

**Supplementary Table 3:** (Top) Dose-response relationship between the intended exposure produced by the irradiator as compared to the dose detected by TLD. (Middle) Dose response relationship between intended exposure from irradiator compared to the dose delivered to a simulated lung -simulated mouse body-encased TLD positioned in the shield aperture. (Bottom) Dose-response relationship between the intended irradiator exposure compared to TLD positioned behind shielding to measure dose delivered to the shielded mouse body.

| **Unshielded Dose-Response Curve** |  |  |  |  |  |  |  |  |
| --- | --- | --- | --- | --- | --- | --- | --- | --- |
| Dose (cGy) | Replicate | Timer (s) | Dose(cGy) | Reading(cGy) | Avg Reading (cGy) | Sigma (%) | Energy Corrected | Energy and Background Corrected |
| 0 | A | 0 | 0 | 0.04 |  |  |  |  |
|  | B |  |  | 0.05 | 0.04 | 17.6 | 0.04 | 0 |
| 50 | A | 40 | 50 | 58.88 |  |  |  |  |
|  | B |  |  | 60.3 | 59.59 | 1.7 | 58.42 | 58.38 |
| 100 | A | 80 | 100.6 | 108.21 |  |  |  |  |
|  | B |  |  | 106.6 | 107.4 | 1.1 | 105.3 | 105.26 |
| 150 | A | 120 | 150.9 | 166.14 |  |  |  |  |
|  | B |  |  | 153.73 | 159.93 | 5.5 | 156.8 | 156.75 |
| 200 | A | 159 | 199 | 224.45 |  |  |  |  |
|  | B |  |  | 223.29 | 223.87 | 0.4 | 219.48 | 219.44 |
| **Shielded Dose-Response Curve in Simulated Mouse Body** |  |  |  |  |  |  |  |  |
| Dose (cGy) | Replicate | Timer (s) | Dose(cGy) | Reading(cGy) | Avg Reading (cGy) | Sigma (%) | Energy Corrected | Energy and Background Corrected |
| **Shield Aperture** |  |  |  |  |  |  |  |  |
| 100 | A | 80 | 100.9 | 75.99 |  |  |  |  |
|  | B |  |  | 74.2 | 75.1 | 1.7 | 73.62 | 73.58 |
| 200 | A | 159 | 199.9 | 156.54 |  |  |  |  |
|  | B |  |  | 152.47 | 154.51 | 1.9 | 151.48 | 151.43 |
| 200 | A | 159 | 199.9 | 151.53 |  |  |  |  |
|  | B |  |  | 148.73 | 150.13 | 1.3 | 147.18 | 147.14 |
| **Shielded** |  |  |  |  |  |  |  |  |
| 100 | A | 80 | 100.9 | 3.46 |  |  |  |  |
|  | B |  |  |  | 3.46 |  | 3.39 | 3.35 |
| 200 | A | 159 | 199.9 | 6.36 |  |  |  |  |
|  | B |  |  | 6.32 | 6.34 | 0.5 | 6.22 | 6.18 |
| 200 | A | 159 | 199.9 | 6.09 |  |  |  |  |
|  | B |  |  | 6.08 | 6.08 | 0.2 | 5.97 | 5.92 |

**Supplementary Figure 1:**
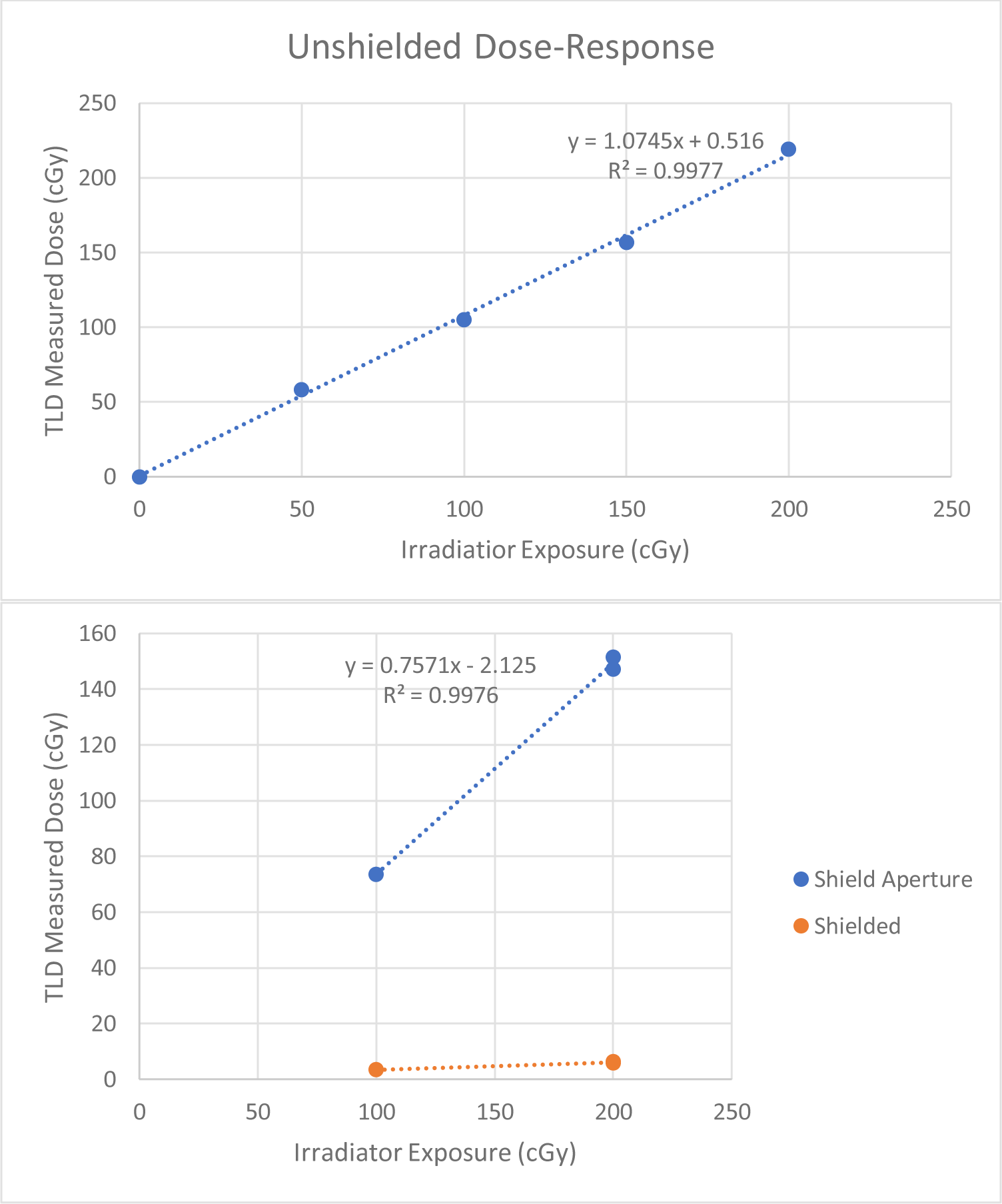
(Top) Dose response relationship between irradiator exposure and TLD measured dose. R2 =0.9977, suggesting a close relationship between intended irradiator exposure and delivered dose to unshielded TLD. (Bottom)

**Supplementary Figure 2:**
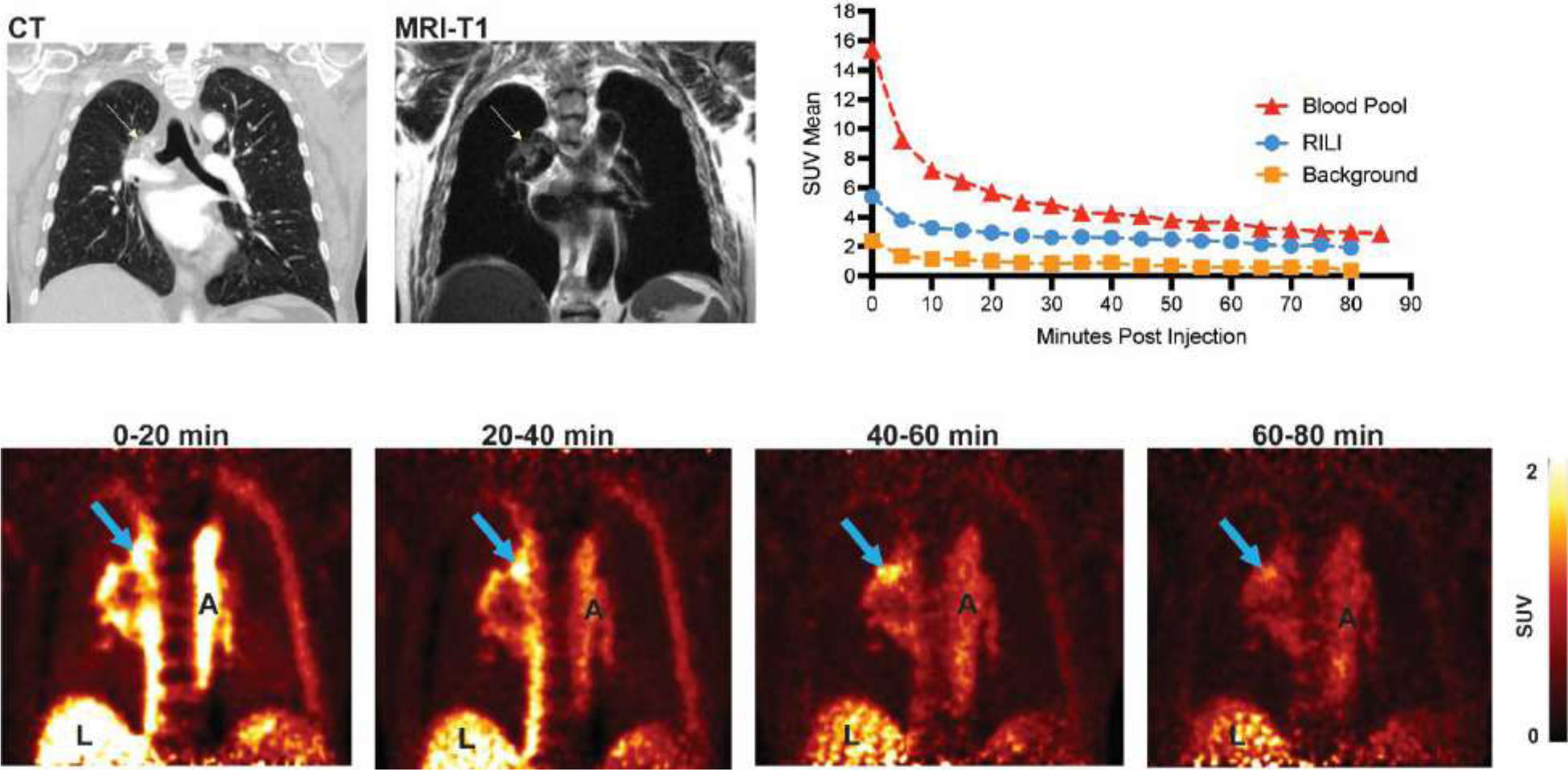
In vivo human imaging. Dynamic imaging of ^68^Ga-CBP8 in a subject with confirmed RILI. (Top row) Coronal HRCT, MR, and PET-MR images. Time activity curves for ^68^Ga-CBP8 in the blood pool, a region of interest of confirmed RILI and for unaffected lung. (Bottom row) Coronal ^68^Ga-CBP8 PET images as a function of time demonstrate probe washout. A- Aorta, L – Liver, Arrow – Area of RILI.

